# Prevalence of diabetes and associated risk factors in Ga Mashie, Accra, Ghana – the CARE Diabetes community-based survey

**DOI:** 10.1101/2024.03.15.24304379

**Authors:** Carlos Salvador Grijalva-Eternod, Kojo Mensah Sedzro, Kafui Adjaye-Gbewonyo, Sandra Boatemaa Kushitor, Swaib Abubaker Lule, Mawuli Komla Kushitor, Akanksha Abhay Marphatia, Ethan Gray, Samuel Amon, Olutobi Adekunle Sanuade, Raphael Baffour Awuah, Leonard Baatiema, Irene Akwo Kretchy, Daniel Arhinful, Kwadwo Ansah Koram, Edward Fottrell

## Abstract

**Background:** Globally, diabetes affects 537 million individuals aged 20-79, significantly undermining their quality of life and economic stability, with the greatest impact in low- and middle-income countries. This study aims to deepen understanding of the diabetes burden in Ga Mashie, an urban-poor area in Accra, Ghana.

**Methods:** We conducted a cluster survey of adults over 25 years in 80 enumeration areas within Ga Mashie, targeting 959 eligible households based on the 2021 census. Household-level data included household membership and structure, water and sanitation, cooking infrastructure, and asset ownership. Individual-level data encompassed demographics, lifestyle behaviours, and biometric measurements. Diabetes was identified through random blood glucose levels ≥11.1 mmol/L or a prior diagnosis, with obesity defined as a body mass index >30 kg/m2 and central obesity as a waist circumference-to-height ratio >0.5. We derived weighted prevalence estimates and compared these estimates by age, sex, and wealth, using unadjusted odds-ratios (OR).

**Results:** The survey, achieving a 67% response rate, covered 854 individuals from 644 households. It unveiled a notable prevalence of non-communicable disease risk factors: 47.2% for alcohol consumption (95% CI: 43.7-50.8), 50.7% for insufficient physical activity (95% CI: 46.0-55.3), 28.9% for unhealthy snack consumption (95% CI: 24.5-33.7), 35.1% for obesity (95% CI: 31.3-39.1), and 74.5% for central obesity (95% CI: 70.8-77.9). Diabetes affected 8.2% of the population aged ≥25 (95% CI: 6.4-10.5), with disparities evident across age, wealth, and sex (2.66 greater odds in females for diabetes [95% CI: 1.38-5.12]).

**Conclusion:** Diabetes and its risk factors are highly prevalent in Ga Mashie, with significant demographic disparities underscoring the need for targeted interventions. The study highlights the critical challenge diabetes poses in urban-poor contexts, emphasizing the necessity for tailored health initiatives to mitigate this burden.

**KEY QUESTIONS:** What is already known on this topic?

- Diabetes and non-communicable diseases (NCDs) present a significant global health challenge, especially in low- and middle-income countries, where there is a notable lack of data on the prevalence and distribution of these conditions and their associated risk factors. The variation in diabetes epidemiology and its risk factors between urban and rural settings highlights the need for context-specific intervention strategies.

What this study adds?

- In Ga Mashie, the prevalence of diabetes is approximately 8%, with over 25% of individuals with diabetes being unaware of their condition and more than a third of those diagnosed not achieving control over the disease. There is also a high prevalence of NCD risk factors, such as insufficient physical activity (73%) and central obesity (75%).
- The distribution of diabetes and NCD risk factors is uneven across different demographic groups, with women and older adults bearing a higher burden of physical inactivity, adiposity, and diagnosed but uncontrolled diabetes, whereas men are more prone to engage in smoking and alcohol consumption.

How might this study affect research, practice, or policy?

- Given the substantial health threat posed by diabetes and NCDs in Ga Mashie, there is a pressing need for interventions at the individual, community, and health system levels. These interventions should be designed with consideration of the unequal disease and risk factor distribution and should aim to address the specific contextual factors driving these disparities.

## INTRODUCTION

Diabetes, a chronic non-communicable disease (NCD) characterised by elevated concentration of blood glucose, poses a significant health challenge, affecting one in ten individuals aged 20-79 years worldwide, approximately 537 million cases (1,2). Among the three primary types of diabetes—type 1, type 2, and gestational diabetes—type 2 diabetes predominates, accounting for over 96% (95% CI: 95.1; 96.8) of cases worldwide (2). The impacts of diabetes are far-reaching, encompassing both micro- and macro-vascular complications such as neuropathy, vision impairment, and stroke. These complications profoundly affect quality of life, longevity, and economic aspects of individuals, households, and communities (3–5).

In the past four decades, diabetes prevalence has exhibited a concerning upward trend, surging from 4.7% in 1980 to 8.5% in 2014 (6). Projections suggest a further rise to 12.2% (783 million cases) by 2045 (7). Low- and middle-income countries (LMICs) are estimated to bear the brunt of this increase, accounting for nearly two-thirds of the projected rise (8). Despite this, half of those living with diabetes from LMICs remain undiagnosed due to a lack of evident warning signs, inconclusive early symptoms, and poor access to screening and diagnosis (7). Although, the burden of diabetes is on the rise in LMICs, urban settings and high-income countries still consistently exhibit higher diabetes prevalence, with regional discrepancies partially attributed to variations in lifestyle and body mass index (BMI) (8). The continent of Africa grapples with a marked rise in diabetes prevalence, from an estimated 3.1% in 1980 to 7.1% in 2014 (9). By 2023, 18.6 million people are projected to be affected by diabetes on the continent (6). Notably, Africa contends with the highest proportion of undiagnosed diabetes cases, with 13 million adults living unaware with this condition (8). This region will experience a significant escalation in diabetes burden, predicted to rise by 156% by 2045, if timely interventions are not implemented (8).

The upward trajectory of diabetes prevalence is intricately tied to urbanization and shifts in dietary and lifestyle patterns (7,8,10). Specific contributing factors include physical inactivity, advancing age, obesity, poor dietary practices, tobacco and alcohol consumption, as well as social and economic determinants such as education, employment, wealth, and social class (11,12). This rising diabetes burden is associated with an increased incidence of both chronic and acute diseases, resulting in compromised quality of life, diminished physical and mental health, premature mortality, heightened healthcare demand, and substantial economic repercussions (10,11,13).

Despite the escalating burden of diabetes, research focusing on the nuanced context of diabetes in most LMICs remains limited (14–16). In Ghana, the prevalence of diabetes is estimated at 6.46% (95% CI: 4.66–8.26%), based on a meta-analysis of studies with varied sampling designs (12). To our knowledge in Accra, Ghana’s urban capital, only two studies have been done to estimate the diabetes prevalence, the last one over a decade ago (17,18). While the role of diet, physical activity, medication, and regular screenings is acknowledged, social, cultural, and economic factors, as well as awareness of one’s diabetic status, significantly influence health behaviours (19). Previous studies conducted in Ghana have identified barriers to diabetes prevention and management, including perceptions of risk, resistance to behaviour change, social stigma, psychosocial burden, economic constraints, medication non-adherence and limited access to healthcare facilities and nutritious foods (20–22). Addressing these challenges necessitates the development of cost-effective diabetes prevention programs tailored to the Ghanaian population. The success of such intervention hinges on an in-depth understanding of local context.

Using data from the ‘Contextual Awareness, Response and Evaluation (CARE): Diabetes in Ghana’ project—a mixed methods study to generate a contextual understanding of diabetes in an urban poor population—this study aimed to generate evidence to further our understanding of the diabetes burden by assessing the prevalence and associated factors from a survey conducted in Ga Mashie, Accra, Ghana.

## MATERIALS AND METHODS

### Ethics

Ethical approval for the CARE project was granted by the Ghana Health Service (GHS-ERC: 017/02/22); Noguchi Memorial Institute for Medical Research Institutional Review Board, University of Ghana (NMIMR-IRB CP 060/21-22); and the Research Ethics Committee at University College London (ID:21541/001). All survey participants provided written informed consent.

### Study design, setting, and population

This study uses data from the CARE survey. A detailed description of the CARE survey methods is available elsewhere (23). Briefly, we undertook the CARE survey in 959 households located in Ga Mashie, Accra, Ghana, between November 10^th^ and December 8^th^, 2022. The locality is an urban poor and densely populated setting, made up of two communities, James Town and Ussher Town. Ga Mashie is inhabited by Indigenous Ga people and migrant populations from across Ghana. Fishing remains a major source of livelihood, though small-scale trading and other commercial activities now dominate the community.

### Sample size estimation

To calculate the sample size, we assumed a diabetes prevalence of 5.0% (12), a precision of 2.0% and a design effect of 2.5. This resulted in an estimated sample of 1,242 individuals. We further assumed an average of two eligible adults per household and a 10% individual refusal to participate. This resulted in an estimated sample of 684 households. However, based on previous field experiences of some of the authors, we further assumed that 40% of households would be found empty or non-traceable, leading to a final estimated sample size of 958 households.

### Survey sampling

We used the 2021 census, conducted by the Ghana Statistical Service (GSS) (24), as our sample frame. To simplify the process and ensure a broad geographical representation across all 80 census enumeration areas (EAs) within Ga Mashie, we requested the GSS to select twelve households from each EA through simple random sampling. This approach yielded in a final sample of 959 households, as one of the EAs had only eleven households randomly sampled.

During the survey, we collected household-level data from all randomly selected households that agreed participation in the survey, and individual-level data from all consenting participants from those households that met the eligibility criteria, which were to be a permanent resident of the household and to be aged ≥25 years. We excluded women who were pregnant or had given birth within the past 6 months. We also excluded anyone deemed unable to provide informed consent or complete the survey such as individuals with impaired mental capacity or who were deaf and unaided. We defined a household as either a single person living alone or a group of people who may not be related but live at the same address and share cooking facilities, a living room, sitting room, or dining area (25). We defined a permanent resident as someone who has lived in the selected household for the past 12 months.

### Patient and Public Involvement

Prior to the survey, we undertook a community engagement activity. This was a participatory event engaging members from different sections of the community, including chiefs, queen mothers (women leaders in the community), market leaders (controllers of specific market produce), fisherfolk, butchers, boxers, and health care providers. The activity was designed to introduce and explain the project, answer questions, and obtain the views of the potential project participants.

### Training of data collectors and quality assurance

Forty enumerators were recruited and provided a five-day training in survey tools and data collection procedures (October 31^st^ – November 4^th^, 2022). The training encompassed obtaining informed consent, conducting participant interviews, ensuring confidentiality, measuring and recording biometric data (such as anthropometry and blood glucose concentration), and utilising Open Data Kit (ODK) questionnaires via Android mobile devices. Enumerators also received training on the detailed Standard Operating Procedures governing fieldwork.

We conducted a pilot survey with 50 households in the La Dade-Kotopon Municipal area of Accra. The pre-test assessed field procedures and data processes, leading to adjustments aimed at enhancing data quality for the main survey. The data from the pilot survey was not included in this analysis.

We encrypted and password-protected all mobile devices, to ensure security and confidentiality. We assigned each household on the sampling list a unique identifier. On arrival to the household, the head of household reviewed and confirmed their identifying information, including structure and house number, as well as address, which was captured in the census. Distinct questionnaires for household- and individual-level data were administered using ODK questionnaires.

Data were regularly uploaded securely to an online server for storage, cleaning, coding, and anonymisation. Data from the server were checked regularly during data collection to identify potential errors. These identified errors were queried, and correction actioned following discussions with the survey supervisors and field teams.

### Data and measurements

We collected data at the household-level from either the household head or the primary household respondent. They provided information on the composition of the household, including the age of each members and their relationship to the household head; the household’s water and sanitation infrastructure, detailing access to private or communal piped water and toilets facilities; cooking infrastructure e.g., the primary type of fuel used for cooking; and ownership of 23 different items. From consenting individuals, we obtained information on age, sex, ethnicity, religion, marital status, highest education level achieved, and whether they were engaged in remunerated work.

We collected data on tobacco and alcohol consumption by utilising relevant items from the World Health Organisation (WHO) STEPS Questionnaire (26). We evaluated individuals’ diets using the 31-item Diet Quality Questionnaire (DQQ), adapted to the Ghanaian context (27). We gauged physical activity using the 16-item Global Physical Activity Questionnaire (GPAQ) (28). We inquired about medical history regarding 12 common NCDs, which included diabetes.

We also collected individual biometric data. We measured and recorded weight, height, and waist circumference. We measured weight to a precision of 0.1 kg, from participants standing on a digital scale (GLC-D-200KG digital body scale, GreenLife Canada) with weight evenly distributed between both feet, arms hanging freely at the sides, and wearing light clothing. We measured height using stadiometers to a precision of 0.1 cm, with participants standing without shoes, feet together, aligning heels, buttocks, and upper back vertically, and orienting the head according to the Frankfurt plane. We measured waist circumference (WC) to a precision of 0.1 cm at the navel level using a measuring tape, while participants breathed normally, at the end of a regular expiration.

Trained laboratory technicians measured Random Blood Glucose (RBG) concentrations using point-of-care glucometers (One Touch select plus, LifeScan Europe GmbH 6300 Zug, Switzerland), obtaining capillary blood drops from the middle finger following a finger prick. We also recorded whether survey participants had previously been diagnosed with diabetes by a health professional; and whether they ever received treatment for diabetes.

### Data handling

To help understand the prevalence of diabetes and NCD risk factors affecting young adults, older adults, and the elderly, we categorised age into three groups: 25-44, 45-64, and ≥65 years. We used Principal Component Analysis of household utilities, structure and asset ownership to generate a household wealth index (29). We categorised the generated household wealth index into tertiles, specifically as ‘most poor’, ‘poor’, and ‘least poor’.

We computed the BMI by dividing weight by height squared, measured in kg/m². Obesity was defined as a BMI ≥30 kg/m². We calculated the waist circumference-to-height ratio (WHR). We defined central obesity as either having a WC >102 cm or >88 cm in males and females, respectively; or as having a WHR >0.5.

A participant was identified as living with diabetes if they reported to have received a prior medical diagnosis or were receiving treatment for diabetes, or if they exhibited RBG values ≥11.1 mmol/L (4). While RBG values are not diagnostic for diabetes, they serve as a valuable tool for assessing diabetes risk in population-based surveys, particularly in situations where obtaining alternative measures such as fasting blood glucose or two-hour glucose tolerance tests is challenging (30). We further categorised those found to be living with diabetes as controlled, uncontrolled, and undiagnosed; if they were previously diagnosed and had normal RBG values, were previously diagnosed but had high RBG values, and were not previously diagnosed but had high RBG values, respectively.

From the DQQ, we used the binary salty or fried snack consumption as a negative indicator of the diet; which ask about consumption of plantain or potato chips, indomie (noodles), or any fried food like yam, potato, atomo, spring rolls, chicken, or fish (27).

From the GPAQ, we used the ‘not meeting WHO Recommendations on physical activity for health’ binary indicator, which assessed the prevalence of respondents that failed to meet the WHO recommendations on physical activity for health, that is, 150 minutes of moderate-intensity physical activity per week, or equivalent (28).

### Statistical analysis

We undertook data analysis using Stata (StataCorp. 2023. Stata Statistical Software: Release 18. College Station, TX: StataCorp LLC). We used the survey commands in Stata (prefix *svy*) in the analysis that accounted for the clustered survey design and the weighing probability of each household or individual. We calculated weight as the ratio of the total number of households or individuals in each enumeration area, as recorded in the 2021 census, to the number of households or individuals surveyed for household- and individual-level data analysis, respectively.

To evaluate the prevalence of diabetes and its known risk factors, we estimated the overall and stratified weighted proportions, stratifying by age, wealth, and sex categories. We then tested for associations using chi-square test. We further explored the relationship between age, wealth and sex with diabetes and its known risk factors, deriving crude odds ratios derived using logistic regression analysis.

## RESULTS

### Participants recruitment flow

Figure 1 presents the survey’s participants flow from the original sample of 959 eligible households randomly selected from 80 EAs. Households that were not found, refused participation, and agreed participation were 31.8% (95% CI: 26.6-37.5), 1.61% (95% CI: 0.83-3.11), and 66.6% (95% CI: 61.1-71.7), respectively. Of a total of 1,007 eligible individuals, which were found in the 644 households that agreed to be surveyed (1.56 eligible individuals/household), 13.5% were absent and 1.8% refused participation in the survey. The 854 surveyed individuals belonged to 629 households from 79 EAs.

**Figure 1.**
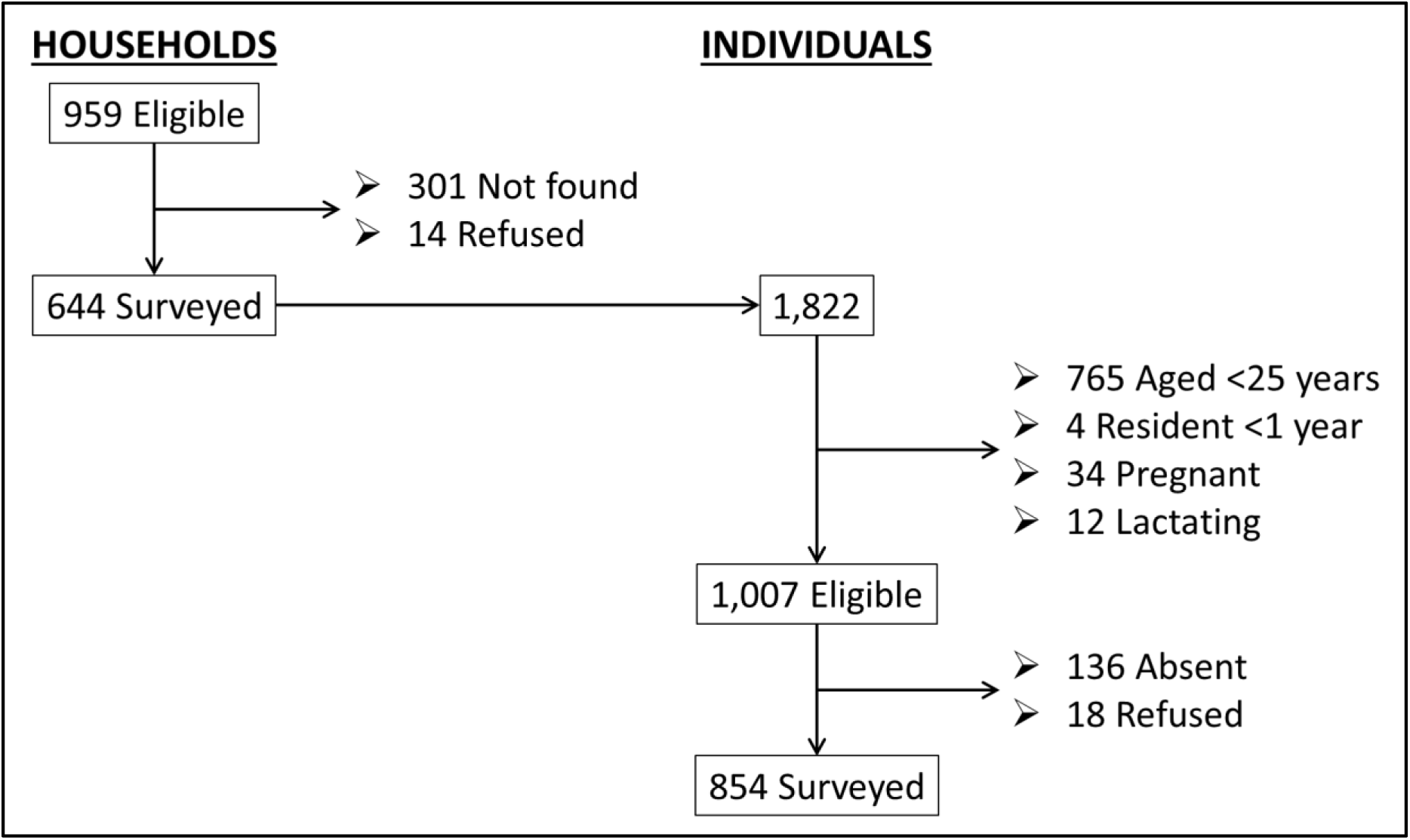
Participants flow.

There appeared to be a bias in the proportion of eligible individuals that were not surveyed because they were absent or refused participation (n=153), as their proportion was greater in the younger age categories (20.4% of those aged 25-44 years vs 3.73% of those aged ≥65 years), in the top wealth tertiles (16.8% of those least poor vs 12.1% of those most poor), and in males (19.9% of males vs 12.3% of females). Nonetheless, this absence did not appear to affect the age, wealth and sex distribution of the surveyed sample compared to the eligible sample (see supplementary **Table S1**).

### Sample characteristics

A table describing the basic characteristics of the 644 surveyed households is provided in the supplementary materials (**Table S2)**. The average participating household comprised of almost three members and slightly over half of these members were females. Over half of the households reported to be female headed, but this proportion showed an inverse relationship with wealth tertiles (64.7% in the most poor households [95% CI: 58.6-70.4] vs 40.0% in the least poor [95% CI: 32.6-47.9]). Most household members were of working age; and wealthier households reported a greater proportion of child dependents (26.4% in the least poor households [95% CI: 22.1-31.3] vs 17.5% in the most poor [95% CI: 13.0-23.3]) but a lower proportion of aged dependents (4.3% in the least poor households [95% CI: 2.6-6.8] vs 10.5% in the most poor [95% CI: 7.8-13.9]). Most households reported shared use of toilet facilities and the fuel used for cooking was different according to wealth tertiles, where households in the most poor tertile used charcoal as their most common fuel (86.9%, 95% CI: 82.1-90.6) and those least poor used gas (57.8%, 95% CI: 49.6-65.6). The characteristics of the surveyed individuals are shown in **Table 1**, stratified by sex, wealth tertiles and age categories. We observed a relationship between sex, wealth, and age; where the most poor tertile had a higher proportion of females, and age showed an inverse relationship with wealth. In Table 1 we also observed other patterns, where ethnic group, religion, marital status, education level, and working status showed significant differences by sex, wealth, and age.

**Table 1.** Individual characteristics.

| Characteristic | Sex |  |  |  | Wealth tertiles |  |  |  |  |  | Age categories (years) |  |  |  |  |  |
| --- | --- | --- | --- | --- | --- | --- | --- | --- | --- | --- | --- | --- | --- | --- | --- | --- |
|  | Male<br>(n=305) |  | Female<br>(n=549) |  | Most poor<br>(n=277) |  | Poor<br>(n=284) |  | Least poor<br>(n=293) |  | 25-44<br>(n=397) |  | 45-64<br>(n=328) |  | 65+<br>(n=129) |  |
|  | % | 95% CI | % | 95% CI | % | 95% CI | % | 95% CI | % | 95% CI | % | 95% CI | % | 95% CI | % | 95% CI |
| Female sex (%) | - | - | - | - | **71.4** | **(65.9,76.4)** | **62.8** | **(55.2,69.8)** | **57.1** | **(51.7,62.3)** | 61.7 | (56.1,67.0) | 63.6 | (58.2,68.8) | 70.1 | (58.2,79.9) |
| Wealth tertiles (%) |  |  |  |  |  |  |  |  |  |  |  |  |  |  |  |  |
| Most poor | **25.9** | **(20.3,32.5)** | **37.0** | **(30.9,43.5)** | - | - | - | - | - | - | **26.0** | **(19.9,33.3)** | **36.1** | **(29.3,43.4)** | **45.1** | **(35.1,55.5)** |
| Poor | **33.1** | **(26.4,40.5)** | **31.9** | **(26.1,38.3)** | - | - | - | - | - | - | **29.5** | **(23.0,37.0)** | **35.8** | **(28.4,43.9)** | **32.8** | **(24.2,42.8)** |
| Least poor | **41.0** | **(34.2,48.1)** | **31.1** | **(25.4,37.4)** | - | - | - | - | - | - | **44.5** | **(37.2,52.0)** | **28.1** | **(21.9,35.3)** | **22.0** | **(14.4,32.2)** |
| Age categories (%) |  |  |  |  |  |  |  |  |  |  |  |  |  |  |  |  |
| 25-44 years | 48.7 | (42.2,55.3) | 44.7 | (40.0,49.5) | **36.6** | **(29.7,44.0)** | **42.0** | **(34.6,49.7)** | **59.1** | **(51.7,66.1)** | - | - | - | - | - | - |
| 45-64 years | 38.7 | (32.0,45.9) | 38.5 | (34.5,42.7) | **42.4** | **(36.8,48.2)** | **42.6** | **(35.0,50.5)** | **31.2** | **(25.4,37.8)** | - | - | - | - | - | - |
| 65+ years | 12.6 | (8.4,18.4) | 16.8 | (13.7,20.3) | **21.0** | **(15.7,27.4)** | **15.4** | **(11.4,20.5)** | **9.7** | **(6.6,13.9)** | - | - | - | - | - | - |
| Ethnic group (%) |  |  |  |  |  |  |  |  |  |  |  |  |  |  |  |  |
| Akan | 13.9 | (10.0,19.0) | 12.1 | (8.7,16.4) | 13.3 | (8.1,20.9) | 13.6 | (9.4,19.2) | 11.4 | (7.4,17.1) | **13.9** | **(10.3,18.5)** | **11.6** | **(7.9,16.8)** | **11.6** | **(6.1,21.0)** |
| Ga-Dangme | 75.8 | (69.7,81.1) | 78.7 | (73.0,83.5) | 80.6 | (72.4,86.7) | 79.1 | (72.7,84.3) | 73.7 | (66.2,79.9) | **72.6** | **(65.9,78.3)** | **81.3** | **(75.6,85.9)** | **84.0** | **(74.7,90.4)** |
| Other | 10.3 | (7.0,14.9) | 9.2 | (6.6,12.7) | 6.2 | (3.3,11.2) | 7.4 | (4.4,12.2) | 14.9 | (11.1,19.7) | **13.5** | **(10.1,18.0)** | **7.1** | **(4.3,11.3)** | **4.3** | **(1.7,10.3)** |
| Religion (%) <sup>1</sup> |  |  |  |  |  |  |  |  |  |  |  |  |  |  |  |  |
| No religion | **8.0** | **(4.9,12.9)** | **3.2** | **(1.8,5.7)** | 8.5 | (4.5,15.5) | 3.7 | (2.0,6.6) | 2.8 | (1.2,6.2) | 5.3 | (3.1,9.1) | 5.7 | (3.2,10.1) | 2.0 | (0.4,9.7) |
| Christian | **60.5** | **(53.6,67.0)** | **70.0** | **(64.3,75.2)** | 67.6 | (60.0,74.4) | 65.3 | (57.3,72.4) | 66.7 | (57.8,74.6) | 61.1 | (54.7,67.1) | 68.4 | (61.6,74.5) | 78.6 | (69.9,85.3) |
| Islam | **16.7** | **(12.2,22.4)** | **10.6** | **(8.2,13.6)** | 9.4 | (6.2,14.0) | 14.1 | (8.3,22.9) | 14.8 | (10.2,21.1) | 16.7 | (12.7,21.7) | 10.0 | (6.3,15.6) | 8.2 | (4.4,15.0) |
| Traditional/Spiritual/Other | **13.9** | **(9.3,20.2)** | **16.1** | **(11.7,21.9)** | 14.0 | (9.7,19.8) | 16.6 | (11.4,23.7) | 15.3 | (8.8,25.3) | 16.4 | (10.9,23.9) | 15.4 | (11.2,20.9) | 11.1 | (6.4,18.5) |
| Currently working (%) | **79.7** | **(73.5,84.8)** | **69.2** | **(64.5,73.6)** | **65.2** | **(59.0,70.9)** | **69.4** | **(63.6,74.7)** | **83.8** | **(77.8,88.5)** | **85.8** | **(81.5,89.2)** | **76.3** | **(70.6,81.2)** | **26.3** | **(19.0,35.2)** |
| Marital status (%) <sup>2</sup> |  |  |  |  |  |  |  |  |  |  |  |  |  |  |  |  |
| Married/Living together | **63.1** | **(56.0,69.7)** | **40.0** | **(35.5,44.6)** | **34.5** | **(28.5,41.0)** | **48.0** | **(39.2,56.9)** | **62.0** | **(53.7,69.6)** | **54.9** | **(49.1,60.6)** | **50.2** | **(42.1,58.3)** | **23.1** | **(15.8,32.7)** |
| Divorced/Separated/Widowed | **17.0** | **(12.7,22.4)** | **42.7** | **(37.8,47.8)** | **45.4** | **(37.9,53.1)** | **34.9** | **(27.7,42.8)** | **20.5** | **(15.5,26.6)** | **11.5** | **(8.3,15.7)** | **43.0** | **(35.9,50.5)** | **75.6** | **(66.2,83.1)** |
| Never married | **19.4** | **(14.6,25.4)** | **17.0** | **(13.0,21.9)** | **19.6** | **(14.1,26.7)** | **16.4** | **(12.2,21.8)** | **17.5** | **(12.2,24.5)** | **33.4** | **(27.6,39.7)** | **6.0** | **(3.2,11.2)** | **1.2** | **(0.2,8.4)** |
| Highest education level (%) |  |  |  |  |  |  |  |  |  |  |  |  |  |  |  |  |
| No education/Pre-school | **9.7** | **(6.4,14.5)** | **14.8** | **(11.2,19.4)** | **19.9** | **(14.5,26.8)** | **13.7** | **(9.3,19.7)** | **5.7** | **(3.0,10.7)** | **6.8** | **(3.9,11.6)** | **13.6** | **(9.6,18.9)** | **29.3** | **(20.0,40.8)** |
| Primary | **15.2** | **(10.3,21.8)** | **21.3** | **(17.3,26.0)** | **28.8** | **(22.3,36.4)** | **18.9** | **(14.7,24.0)** | **10.0** | **(6.2,15.6)** | **17.1** | **(12.6,22.8)** | **24.7** | **(18.7,31.9)** | **11.4** | **(6.7,19.0)** |
| Middle/JHS | **35.2** | **(29.2,41.7)** | **43.1** | **(38.5,47.8)** | **37.4** | **(29.9,45.5)** | **45.7** | **(41.0,50.3)** | **37.8** | **(31.8,44.2)** | **35.6** | **(30.8,40.8)** | **47.9** | **(41.0,55.0)** | **34.1** | **(25.9,43.3)** |
| Secondary/SHS/Higher | **39.9** | **(33.2,47.0)** | **20.8** | **(16.2,26.3)** | **13.9** | **(9.3,20.1)** | **21.8** | **(17.2,27.2)** | **46.4** | **(39.6,53.4)** | **40.5** | **(34.4,46.8)** | **13.8** | **(10.0,18.7)** | **25.1** | **(17.6,34.6)** |

<sup>1</sup> A total of 0.9% (95% CI:0.3,3.0) refused to answer this question. <sup>2</sup> A total of 0.4% (95% CI:0.1,2.9) refused to answer this question.
We used chi-square to test for associations. Bold numbers represent a significant association(p<0.05).

### NCDs behavioural risk factors

A detailed examination of the complex interplay of sex, age, and socioeconomic status in influencing behavioural risk factors for NCDs is evidenced by the data presented in **Table 2**. In our analysis of the sex differences in behavioural risk factors for NCDs as depicted in Table 2, distinct patterns emerged. Women were found to have significantly lower odds of consuming tobacco and alcohol compared to men, with odds ratios of 0.16 [95% CI: 0.07-0.36] and 0.54 [95% CI: 0.38-0.76], respectively. However, women were more likely to not meet the WHO’s physical activity recommendations, with an odds ratio of 3.72 [95% CI: 2.67-5.18]. The consumption of salty or fried snacks did not show significant sex differences, indicating a similar dietary behaviour among both sexes.

**Table 2.** Prevalence and crude ORs of behavioural non-communicable disease risk factors.

|  | Consumed tobacco |  |  |  | Consumed alcohol |  |  |  | Consumed salty or fried snacks <sup>1</sup> |  |  |  | Insufficient physical activity <sup>2</sup> |  |  |  |
| --- | --- | --- | --- | --- | --- | --- | --- | --- | --- | --- | --- | --- | --- | --- | --- | --- |
|  | % | 95% CI | OR | 95% CI | % | 95% CI | OR | 95% CI | % | 95% CI | OR | 95% CI | % | 95% CI | OR | 95% CI |
| Total | 5.5 | (3.9,7.7) | - | - | 47.2 | (43.7,50.8) | - | - | 28.9 | (24.5,33.7) | - | - | 50.7 | [46.0,55.3] | - | - |
| Sex |  |  |  |  |  |  |  |  |  |  |  |  |  |  |  |  |
| Males (n=305) | 11.5 | (7.9,16.4) | ref | - | 57.0 | (50.6,63.2) | ref | - | 29.2 | (23.5,35.6) | ref | - | 30.4 | [24.4,37.2] | ref | - |
| Females (n=549) | 2.1 | (1.0,4.1) | **0.16** | **(0.07,0.36)** | 41.7 | (37.1,46.4) | **0.54** | **(0.38,0.76)** | 28.7 | (23.4,34.6) | 0.98 | (0.68,1.40) | 61.9 | [56.1,67.5] | **3.72** | **[2.67,5.18]** |
| Wealth tertile |  |  |  |  |  |  |  |  |  |  |  |  |  |  |  |  |
| Most poor (n=277) | 7.3 | (4.3,12.2) | ref | - | 46.8 | (40.4,53.2) | ref | - | 26.3 | (19.6,34.4) | ref | - | 63.4 | [55.3,70.7] | ref | - |
| Poor (n=284) | 5.8 | (3.4,9.8) | 0.78 | (0.35,1.76) | 48.5 | (41.6,55.4) | 1.07 | (0.71,1.61) | 27.3 | (21.4,34.0) | 1.05 | (0.64,1.71) | 48.2 | [41.0,55.4] | **0.54** | **[0.35,0.83]** |
| Least poor (n=293) | 3.4 | (1.7,6.8) | 0.45 | (0.18,1.15) | 46.5 | (41.1,52.0) | 0.99 | (0.72,1.36) | 32.8 | (26.6,39.6) | 1.36 | (0.90,2.07) | 41.1 | [34.0,48.5] | **0.40** | **[0.26,0.62]** |
| Age categories |  |  |  |  |  |  |  |  |  |  |  |  |  |  |  |  |
| 25-44 years (n=397) | 7.5 | (4.6,11.9) | ref | - | 49.7 | (45.1,54.4) | ref | - | 33.5 | (27.2,40.5) | ref | - | 38.1 | [32.9,43.5] | ref | - |
| 45-64 years (n=328) | 4.1 | (2.2,7.7) | 0.53 | (0.22,1.28) | 48.6 | (42.6,54.6) | 0.96 | (0.70,1.30) | 25.2 | (19.7,31.6) | **0.67** | **(0.46,0.97)** | 52.9 | [46.1,59.6] | **1.83** | **[1.37,2.44]** |
| 65+ years (n=129) | 2.7 | (0.7,9.7) | 0.35 | (0.08,1.48) | 35.4 | (25.3,47.0) | **0.55** | **(0.33,0.92)** | 24.7 | (16.3,35.7) | 0.65 | (0.36,1.19) | 81.5 | [72.1,88.2] | **7.17** | **[4.16,12.3]** |

<sup>1</sup> From the DQQ, we used the binary salty or fried snack consumption as a negative indicator of the diet. <sup>2</sup> From the GPAQ, we used the 'not meeting WHO Recommendations on physical activity for health' binary indicator.
We used logistic regression to assess odds ratios. Bold numbers represent a significant association (p<0.05).

Age-related trends further delineated risk behaviours, with older participants (45-64 years) showing reduced odds of alcohol consumption and intake of salty or fried snacks compared to their younger counterparts, as outlined in Table 2. A clear age-associated increase in the odds of not fulfilling WHO’s physical activity guidelines was observed, especially pronounced among older age groups, highlighting a trend towards decreased physical activity with aging.

Moreover, our findings indicate that socioeconomic status, represented by wealth tertiles, was not significantly associated with NCD risk factors, with the notable exception of physical activity. Here, individuals in the two higher wealth tertiles displayed lower odds of failing to meet the WHO physical activity recommendations compared to those in the most impoverished group, suggesting that economic factors may play a role in physical activity levels but not in other behavioural risk factors.

### Diabetes prevalence and NCD metabolic risk factors

**Table 3** delineates the prevalence and odds ratios for diabetes within the Ga Mashie population. The findings reveal a diabetes prevalence of 8.2% among individuals (95% CI: 6.4-10.5) and 11.5% at the household level, indicating at least one member living with diabetes (95% CI: 9.1-14.4). Notably, a significant gender disparity is observed, with females exhibiting a higher diabetes prevalence by 6.3 percentage points (95% CI: 2.7-9.9) and a markedly increased odds ratio of 3.59 (95% CI: 1.55-8.30) when compared to males.

**Table 3.** Prevalence and crude ORs of diabetes and the metabolic risk factors obesity and central obesity.

|  | Diabetes <sup>1</sup> |  |  |  | Obesity (BMI >30kg/m<sup>2</sup>) |  |  |  | Central Obesity (WHR >0.5) |  |  |  |
| --- | --- | --- | --- | --- | --- | --- | --- | --- | --- | --- | --- | --- |
|  | % | 95% CI | OR | 95% CI | % | 95% CI | OR | 95% CI | % | 95% CI | OR | 95% CI |
| Total | 8.2 | (6.4,10.5) | - | - | 35.1 | (31.3,39.1) | - | - | 74.5 | (70.8,77.9) | - | - |
| Sex |  |  |  |  |  |  |  |  |  |  |  |  |
| Males (n=305) | 4.2 | (2.4,7.4) | ref | - | 13.8 | (9.0,20.5) | ref | - | 55.3 | (49.1,61.4) | ref | - |
| Females (n=545) | 10.5 | (8.0,13.7) | **2.66** | **(1.38,5.12)** | 47.4 | (42.5,52.3) | **5.63** | **(3.40,9.31)** | 85.5 | (81.5,88.8) | **4.77** | **(3.36,6.79)** |
| Wealth tertile |  |  |  |  |  |  |  |  |  |  |  |  |
| Most poor (n=277) | 9.3 | (6.4,13.2) | ref | - | 36.9 | (29.9,44.5) | ref | - | 73.6 | (67.9,78.7) | ref | - |
| Poor (n=283) | 10.0 | (6.6,14.8) | 1.08 | (0.61,1.93) | 33.5 | (27.9,39.7) | 0.86 | (0.56,1.32) | 72.7 | (64.8,79.5) | 0.96 | (0.63,1.45) |
| Least poor (n=290) | 5.6 | (3.3,9.3) | 0.58 | (0.29,1.15) | 35.0 | (29.5,40.9) | 0.92 | (0.62,1.38) | 77.0 | (71.7,81.6) | 1.20 | (0.80,1.80) |
| Age category |  |  |  |  |  |  |  |  |  |  |  |  |
| 25-44 years (n=395) | 0.6 | (0.2,2.4) | ref | - | 29.5 | (25.1,34.2) | ref | - | 64.8 | (59.9,69.5) | ref | - |
| 45-64 years (n=327) | 14.4 | (10.1,20.1) | **26.8** | **(6.40,112.5)** | 44.5 | (38.4,50.8) | **1.92** | **(1.37,2.68)** | 83.2 | (76.6,88.3) | **2.69** | **(1.69,4.3)** |
| 65+ years (n=128) | 15.8 | (9.9,24.1) | **29.9** | **(6.77,131.9)** | 28.7 | (19.6,39.9) | 0.96 | (0.57,1.61) | 81.4 | (72.7,87.8) | **2.38** | **(1.43,3.96)** |

BMI: Body mass index. WC: Waist circumference. WHR: Waist-to-height ratio.
<sup>1</sup> A person was defined as living with diabetes if they reported to have received a prior medical diagnosis or were receiving treatment for diabetes, or if they exhibited RBG values $\geq 11.1$ mmol/L . We used logistic regression to assess odds ratios. Bold numbers represent a significant association ( $p < 0.05$ )

Further analysis indicates that older age groups possess significantly higher diabetes prevalence and odds ratios in comparison to the younger cohort, highlighting age as a critical factor in diabetes risk. However, the distribution of diabetes prevalence appears uniform across various wealth tertiles, suggesting economic status does not significantly influence diabetes risk in this community. Additionally, Table 3 presents obesity estimates, showing females with significantly higher odds of both obesity and central obesity. Age-related differences are pronounced in central obesity across all older groups, whereas only the middle-aged group (45-64 years) demonstrates increased odds for obesity.

The prevalence of controlled, uncontrolled, and undiagnosed diabetes in Ga Mashie was 3.21% (95% CI: 2.17, 4.70), 2.78% (95% CI: 1.85, 4.16), and 2.24% (95% CI: 1.41, 3.54), respectively; and they represented 39.0% (95% CI: 29.1, 49.9), 33.8% (95% CI: 23.9, 45.4), and 27.2% (95% CI: 17.3, 40.2) of the totality of the diabetes prevalence, respectively. Figure 2 presents a visual distribution of the values of RBG by sex and age, where we can visually compare controlled, uncontrolled, and undiagnosed diabetes. **Table 4** presents an analysis of controlled, uncontrolled, and undiagnosed diabetes status, by sex, wealth, and age -relative to the total prevalence. For sex, we observed that females presented with a greater prevalence (a 28-percentage points difference, 95% CI: 8.3; 47.7) and greater odds of uncontrolled diabetes, when compared to men. The opposite pattern was observed in females for controlled diabetes. Females presented also with lower odds of undiagnosed diabetes.

**Figure 2.**
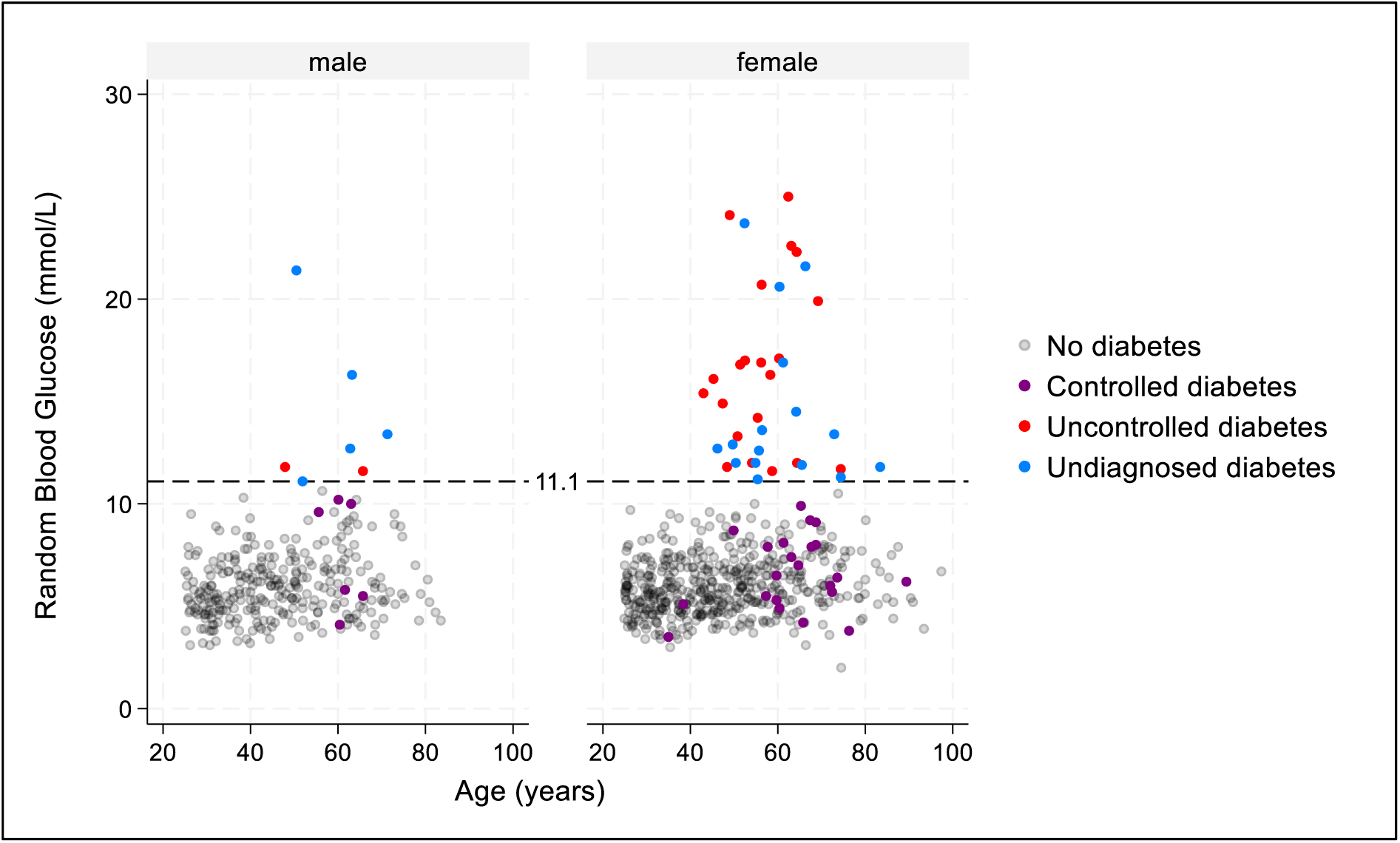
Random blood glucose concentration against age, by sex and diabetes status.

**Table 4.** Relative prevalence and crude odds ratios for controlled, uncontrolled, and undiagnosed diabetes.

|  | Controlled diabetes<br>(Previous diagnosis, but normal RBG) |  |  |  | Uncontrolled diabetes<br>(Previous diagnosis, and high RBG) |  |  |  | Undiagnosed diabetes<br>(No previous diagnosis, but high RBG) |  |  |  |
| --- | --- | --- | --- | --- | --- | --- | --- | --- | --- | --- | --- | --- |
|  | % | 95% CI | OR | 95% CI | % | 95% CI | OR | 95% CI | % | 95% CI | OR | 95% CI |
| All | 39.0 | (29.1, 49.9) | - | - | 33.8 | (23.9, 45.4) | - | - | 27.2 | (17.3; 40.2) | - | - |
| Sex |  |  |  |  |  |  |  |  |  |  |  |  |
| Male | 45.2 | (20.3,72.8) | ref | - | 11.0 | (2.7,35.6) | ref | - | 43.8 | (17.9,73.5) | ref | - |
| Female | 37.6 | (26.9,49.6) | **0.73** | **(0.69,0.77)** | 39.0 | (27.7,51.7) | **5.17** | **(4.79,5.57)** | 23.4 | (14.0,36.5) | **0.39** | **(0.37,0.42)** |
| Wealth tertile |  |  |  |  |  |  |  |  |  |  |  |  |
| Most poor | 20.4 | (8.85,40.5) | ref | - | 42.3 | (22.3,65.2) | ref | - | 37.3 | (19.3,59.6) | ref | - |
| Poor | 44.6 | (24.9,66.1) | **3.13** | **(2.94,3.34)** | 24.4 | (10.7,46.7) | **0.44** | **(0.42,0.47)** | 31.0 | (15.9,51.4) | **0.75** | **(0.71,0.80)** |
| Least poor | 59.0 | (36.4,78.3) | **5.60** | **(5.25,5.96)** | 35.9 | (17.7,59.3) | **0.76** | **(0.72,0.81)** | 5.11 | (0.63,31.5) | **0.09** | **(0.08,0.10)** |
| Age category |  |  |  |  |  |  |  |  |  |  |  |  |
| 25-44 years | 36.3 | (28.7,4.46) | ref | - | 63.7 | (28.7,12.6) | ref | - | 0.00 | - | - | - |
| 45-64 years | 29.4 | (6.57,18.0) | **0.73** | **(0.69,0.77)** | 39.9 | (6.09,28.4) | **0.38** | **(0.36,0.40)** | 30.8 | (7.50,17.9) | ref | - |
| 65+ years | 61.5 | (11.4,37.8) | **2.81** | **(2.65,2.97)** | 16.1 | (10.1,4.08) | **0.11** | **(0.10,0.12)** | 22.3 | (9.20,8.98) | **0.65** | **(0.61,0.69)** |

RBG: Random blood glucose.
We used logistic regression to assess odds ratios. Bold numbers represent a significant association (p<0.05)

Socioeconomic status was also significantly associated with diabetes status. For instance, individuals in the highest socioeconomic status had significantly lower odds of uncontrolled diabetes (OR = 0.76, 95% CI: 0.72, 0.81), and greater odds of controlled diabetes (OR = 5.60, 95% CI: 5.25, 5.96), when compared to the lowest socioeconomic status. This same socioeconomic group presented very low odds of having undiagnosed diabetes.

Age emerged as another significant correlate of diabetes status, with individuals aged 65 and above displaying greater odds of controlled diabetes (OR = 2.81, 95% CI: 2.65, 2.97), and lower odds of uncontrolled or undiagnosed diabetes (OR = 0.11 (95% CI: 0.10, 0.12) and OR = 0.65 (95% CI: 0.61, 0.69) respectively).

## DISCUSSION

### Main findings

This study marks an important advancement in understanding diabetes prevalence in the Ga Mashie locality, an urban-poor enclave in Accra. The survey revealed a significant prevalence of diabetes, affecting over one in ten households within this community. Additionally, it showed the extensive presence of various NCD risk factors among the population. Notably, our results uncovered clear disparities in the prevalence of diabetes and NCD risk factors across different demographic groups. Age and wealth status were identified as key factors, highlighting the varied impact on distinct population segments. Particularly striking was the gender disparity observed, where females were found to carry a disproportionately higher burden of diabetes and numerous NCD risk factors. The study also underscored the increased likelihood of females experiencing uncontrolled diabetes.

### Diabetes prevalence

In this study we revealed a significantly higher prevalence of diabetes within the Ga Mashie area compared to previous regional studies conducted in 2002 by Amoah et al. (17) and in 2011 by Vuvor et al. (18). Amoah et al. reported a diabetes prevalence of 6.3% (95% CI: 5.61, 6.99) in Greater Accra, while Vuvor et al. documented a prevalence of 3.9% (95% CI: 2.3, 5.5) in peri-urban Accra. It is important to note that both earlier studies utilized fasting blood glucose determination, and Amoah et al. a 2-hour glucose tolerance test, different from our methods. These methodological differences, known to impact prevalence rates (31), alongside variations in the sampling universe, might contribute to the disparities observed. Despite these methodological distinctions, our study’s identification of a notably higher prevalence aligns with projections indicating a rising trend of diabetes in the region (8).

### NCD risk factors

The prevalence of alcohol and tobacco consumption in this community was high and we observed a sex-difference, with men reporting a higher consumption in both. In this context, there are factors, perceived by the community, which are potentially driving alcohol consumption and smoking in this region. These factors include economic hardship (e.g., unemployment), psycho-social hardships and anxieties, illness management (e.g., pain suppression), and socio-cultural beliefs (e.g., aphrodisiac, magic) (32). For instance, funerals are local social and important events that facilitate alcohol consumption (32).

Eating habits associated with NCDs, i.e., consumption of salty or fried snacks, were also highly prevalent in this context, suggesting the need for action to improve eating behaviours. However, this call to action in this context is not without challenges. Mensah et al. found that a greater consumption of fruits and vegetables was perceived by the community to be determined by availability, cost, personal preferences, and knowledge and belief (32); with personal preferences and cost considered the main barriers to a higher consumption. This is supported by the work of Suglo et al. which found that people are knowledgeable of what constitutes a healthy diet and what foods are suitable for people living with diabetes in Ga Mashie (33); nonetheless, a healthy diet was perceived as expensive, with cost as a major barrier to consumption of healthy food (33). Furthermore, the adherence to self-care behaviours among people living with diabetes is also a challenge as demonstrated by Opoku et al., where the mean number of days per week adherence to dietary recommendations was 3.9-4.4 days/week; and where only 2.7% of people living with diabetes adhere daily to these recommendations (34).

Insufficient physical activity in this context was found to be highly prevalent, with women presenting with a significantly higher burden, i.e., 62% of women do not engage in sufficient physical activity, compared to 30% of men. Evidence of this gender gap can be explained by gender norms affecting physical activity, as shown by the work of Amin et al. (35), where reported socio-structural barriers for physical activity included concerns about social ridicule or embarrassment, safety during outdoor activities, a lack of culturally appropriate exercise facilities, and high social and work demands. For people living with diabetes, chronic illness-related factors also hindered their exercise participation. Nonetheless, evidence suggests that physical activity interventions in this context might be feasible, and could lead to significant improvement of glycaemic control and reduction of NCD risk factors like waist circumference (35,36).

### Sex-difference in the burden of diabetes and NCD risk factors

In this context we observed that women had a higher burden of diabetes and some NCD risk factors such as obesity and insufficient physical activity. Conversely, men presented a higher risk of alcohol and tobacco consumption. In this context, obesity is understood as a sign of good living and good health and men are understood to find larger women attractive; which might motivate some women to desire to be overweight (37). Weight management is mainly determined by social representation of being fat or slim (37,38).

Contrary to our findings, a recent study that included data from five West African countries found that diabetes and NCD risk factors burden did not vary by sex (39). However, another study also found that in clinical settings, a larger proportion of people living with diabetes are women (40). In our study, although proportionately more eligible men were not surveyed, it is improbable that this absence significantly impacted our results, as the age, sex, and wealth distributions observed in both the eligible and surveyed populations remained similar (see Table S1) (41).

### Uncontrolled diabetes

Among people living with diabetes, a large proportion presented with uncontrolled diabetes defined as having a RBG values ≥11.1mmol/l and a prior diagnosis of diabetes. Uncontrolled diabetes affected women disproportionately – a finding observed elsewhere in Accra and reportedly exacerbated during the COVID-19 era (42).

Consistent self-care among individuals living with diabetes presents notable challenges, reflecting varied adherence levels to medication and care recommendations (34). Barriers to effective self-management are likely to include limited knowledge about diabetes dietary strategies, financial constraints, treatment non-compliance, restricted access to glucometers, inadequate social support, and prolonged waiting times at healthcare facilities (43). Stigma is also a known challenge in the control of diabetes (44). People living with diabetes in other settings have reported that due to stigma, they often keep their diagnoses to themselves and this is linked with non-adherence to treatment and self-management (45). The impact of stigma might be greater among women as women living with diabetes reported a greater risk of divorce if their partners knew about their diagnosis (45). These insights underscore the importance of improved education about diabetes and dietary management strategies, as well as the pivotal role of robust social support from both community networks and organizations in addressing these multifaceted challenges.

Availability, use, and the poor regulation of pluralistic health centres for care is pervasive in the Ga Mashie community and this might also contribute to the manifestation of uncontrolled diabetes (46). Medical care often begins as a combination of allopathic and alternative medicine care, which then shifts fully to other forms of treatment, often this shift being influenced by perceived cheaper costs (46).

Our finding that women in the Ga Mashie present with greater risk of uncontrolled diabetes has also been observed elsewhere in Accra. Women were found to have an increased likelihood of poor glycaemic control; and the prevalence of this poor control was exacerbated during the COVID-19 era (42).

### Undiagnosed diabetes

Pregnant women represent the highest number of healthcare users globally and in Ghana 88% of antenatal care facilities are reported to screen for gestational diabetes (47), potentially explaining why a greater proportion of men present with undiagnosed diabetes in this context. It is also possible that a low relational continuity of care, defined as an ongoing personal relationship between care providers and patients guided by personal trust and a sense of responsibility (48), could partly explain this greater proportion in men, as they often present lower continuity of care (44). In addition, self-denial due to stigma and perceived high cost of treatment, may also contribute to this observed phenomenon (49).

### Limitations and Strengths

There were limitations to our study. We surveyed a smaller sample than originally anticipated based on our assumptions. This might have given us a greater pixelation in our observed patterns of disease and may account for some of the potential contradictions with our findings and that of others, as described above. We use RBG as one of the criteria for defining diabetes, rather than fasting blood glucose or a 2-hour glucose tolerance test (31), although other studies have confirmed the value of RBG measurement for assessing diabetes prevalence and mortality risk in populations (30,50,51). Lastly, in our survey a greater proportion of men were missing and not surveyed (Table S1). Nonetheless, we think this is unlikely to affect the pattern of results observed, as the sex, age, and wealth proportions observed in the surveyed and non-surveyed population were comparable. Our study also has methodological strengths. Our survey design and sampling methodology is robust and representative allowing us to draft inferences from the population of interest in Ga Mashie area. Rigorous training and pilot testing, robust data collection procedures, field supervision and data quality control measures using the most up-to-date digital systems are likely to have increased the quality of the data obtained from this population.

## Conclusion

Diabetes and NCD risk factors are highly prevalent in the Ga Mashie, and they appear to have increased in the past decade. There are inequities in these burdens by age, wealth, and importantly, sex. These nuanced disparities underscore the need for targeted and inclusive interventions that account for multifaceted variations within this community. Further investigation is warranted to better understand the drivers and implications of the escalating diabetes burden in the Ga Mashie area.

## Data Availability

All data produced in the present study are available upon reasonable request to the authors.

## ACKNOWLEDGEMENTS

The authors thank Lucy Twumwaah from the Ghana Statistical Services for conducting the sampling and provision of the map of the study area, as well as the Ga Mashie Development Authority (GAMADA) for the support offered during the survey preparation. The authors also thank the support of the CARE Diabetes Team members Moses Aikins, Publa Antwi, Vida Asah-Ayeh, Ann Blandford, Ama de-Graft Aikins, Hassan Haghparast-Bidgoli, Sarah Hawkes, Hannah Jennings, Ernestina Korleki Dankyi, Rolando Leiva-Granados, Lydia Okoibhole, Emeline Rougeaux, Daniel Strachan, Megan Vaughan, Haim Yacobi.

## DECLARATION OF INTEREST

None

## FUNDING

The project is supported by funds from the Medical Research Council (MRC) through the United Kingdom Research and Innovation (UKRI), grant number MR/T029919/1. The funder has no role in study design and conduct of this study.

## CONTRIBUTIONS

Conception or design of the work: CSG-E, EF, IAK, KA-G, KK, KMS, SBK.

Acquisition of data: CSG-E, DA, IAK, KMS, MKK, OAS, SA, SAL, SBK.

Analysis, or interpretation of data: AAM, CSG-E, EF, EG, IAK, KA-G, KK, KMS, LB, MKK, OS, SAL, SBK.

Drafted the work: CSG-E.

Revised it critically for important intellectual content: All authors.

Approved the version to be published: All authors.

Agree to submit: All authors.

## SUPPLEMENTARY MATERIALS

**Table S1.** Age, wealth, and sex of eligible and surveyed household members.

|  | Eligible |  | Surveyed |  | Non-surveyed* |
| --- | --- | --- | --- | --- | --- |
| Sex | N | % (95% CI) | n | % (95% CI) | n/N (%) |
| Male | 381 | 38.7 (36.0; 41.6) | 305 | 36.3 (33.2; 39.6) | 76/381 (19.9%) |
| Female | 626 | 61.3 (58.5; 64.0) | 549 | 63.7 (60.4; 66.8) | 77/626 (12.3%) |
| Wealth tertile | n | % (95% CI) | n | % (95% CI) | n (%) |
| Most poor | 315 | 30.6 (24.9; 37.0) | 277 | 33.0 (27.9; 38.5) | 30/315 (12.1%) |
| Poor | 340 | 33.0 (27.7; 38.9) | 284 | 32.3 (27.3; 37.8) | 56/340 (16.5%) |
| Least Poor | 352 | 36.4 (31.1; 42.0) | 293 | 34.7 (29.6; 40.2) | 59/352 (16.8%) |
| Age category | n | % (95% CI) | n | % (95% CI) | n (%) |
| 25-44 years | 499 | 49.2 (45.7; 52.7) | 397 | 46.2 (42.3; 50.1) | 102/499 (20.4%) |
| 45-64 years | 374 | 37.6 (34.6; 40.6) | 328 | 38.6 (35.2; 42.1) | 46/374 (12.3%) |
| ≥65 years | 134 | 13.3 (10.9; 16.0) | 129 | 15.3 (13.0; 17.9) | 5/134 (3.73%) |
| Total | 1,007 | 100% | 854 | 100% | 153/1,007 (15.2%) |

\* The percentage of non-surveyed household members is in relation to the eligible sample.

**Table S2.** Household characteristics.

| Characteristic | Total (n=644) |  | Wealth Tertiles |  |  |  |  |  |
| --- | --- | --- | --- | --- | --- | --- | --- | --- |
|  |  |  | Most poor (n=215) |  | Poor (n=215) |  | Least poor (n=214) |  |
|  | % or mean | 95% CI | % or mean | 95% CI | % or mean | 95% CI | % or mean | 95% CI |
| Household size (members) | 2.84 | (2.76, 2.92) | 2.51 | (2.41, 2.61) | 2.92 | (2.79, 3.05) | 3.06 | (2.93, 3.19) |
| Female headed household (%) | 52.9 | (48.7, 57.0) | **64.7** | **(58.6, 70.4)** | **54.0** | **(46.4, 61.4)** | **40.0** | **(32.6, 47.9)** |
| Female member (%) | 57.9 | (55.3, 60.4) | 58.9 | (54.8, 62.8) | 59.8 | (55.5, 64.0) | 55.3 | (50.7, 59.9) |
| Family structure (%) |  |  |  |  |  |  |  |  |
| child dependent (<15 years) | 23.4 | (21.0, 26.1) | **17.5** | **(13.0, 23.3)** | **25.2** | **(22.0, 28.7)** | **26.4** | **(22.1, 31.3)** |
| working age adult (15-64 years) | 69.2 | (66.8, 71.6) | **72.0** | **(66.6, 76.8)** | **66.8** | **(63.1, 70.3)** | **69.3** | **(64.5, 73.7)** |
| aged dependent (≥65 years) | 7.30 | (5.9, 9.0) | **10.5** | **(7.8, 13.9)** | **8.00** | **(6.0, 10.5)** | **4.30** | **(2.6, 6.8)** |
| Household with a pregnant woman (%) | 5.70 | (4.4, 7.4) | **8.1** | **(5.4, 12.0)** | **2.6** | **(1.3, 4.9)** | **6.7** | **(4.6, 9.7)** |
| Shared toilet with other households (%) | 89.6 | (86.4, 92.2) | 94.3 | (89.6, 96.9) | 92.7 | (87.1, 95.9) | 83.1 | (75.5, 88.7) |
| Fuel for Cooking (%) |  |  |  |  |  |  |  |  |
| Charcoal | 65.7 | (61.4, 69.8) | **86.9** | **(82.1, 90.6)** | **74.0** | **(66.3, 80.5)** | **37.5** | **(29.9, 45.8)** |
| LPG | 29.6 | (25.8, 33.7) | **8.6** | **(5.4, 13.5)** | **21.1** | **(15.1, 28.5)** | **57.8** | **(49.6, 65.6)** |

We used chi-square to test for associations. Bold numbers represent a significant association ( $p < 0.05$ ).

## Notes

### Competing Interest Statement

The authors have declared no competing interest.

### Author Declarations

Ethical approval for the CARE project was granted by the Ghana Health Service (GHS-ERC: 017/02/22); Noguchi Memorial Institute for Medical Research Institutional Review Board, University of Ghana (NMIMR-IRB CP 060/21-22); and the Research Ethics Committee at University College London (ID:21541/001).

### Summary of Updates

In this new version we have updated the values of the indicator for insufficient physical activity, which is now estimated to be 50.7% (95% CI: 46.0-55.3). We have also included an author, Raphael Baffour Awuah.

